# Large-scale implementation of pooled RNA-extraction and RT-PCR for SARS-CoV-2 detection

**DOI:** 10.1101/2020.04.17.20069062

**Authors:** Roni Ben-Ami, Agnes Klochendler, Matan Seidel, Tal Sido, Ori Gurel-Gurevich, Moran Yassour, Eran Meshorer, Gil Benedek, Irit Fogel, Esther Oiknine-Djian, Asaf Gertler, Zeev Rotstein, Bruno Lavi, Yuval Dor, Dana G. Wolf, Maayan Salton, Yotam Drier, The Hebrew University-Hadassah COVID-19 diagnosis team

## Abstract

Testing for active SARS-CoV-2 infection is a fundamental tool in the public health measures taken to control the COVID-19 pandemic. Due to the overwhelming use of SARS-CoV-2 RT-PCR tests worldwide, availability of test kits has become a major bottleneck. Here we demonstrate pooling strategies to perform RNA extraction and RT-PCR in pools, significantly increasing throughput while maintaining clinical sensitivity. We implemented the method in a routine clinical diagnosis setting of asymptomatic populations, and already tested 5,464 individuals for SARS-CoV-2 using 731 RNA extraction and RT-PCR kits. We identified six SARS-CoV-2 positive patients corresponding to 0.11% of the tested population.

## Introduction

An emerging novel severe acute respiratory syndrome-related coronavirus, SARS-CoV-2, is the virus behind the global COVID-19 pandemic. Among the foremost priorities to facilitate efficient public health interventions is a reliable and accessible diagnosis of an active SARS-CoV-2 infection. The standard laboratory diagnosis of COVID-19 involves three main steps, namely, viral inactivation and lysis of the nasopharyngeal swab sample, extraction (or purification) of viral RNA, and reverse transcription (RT)-PCR. Due to the rapid spread of the virus and the increasing demand for tests, the limited availability of test reagents, mainly RNA extraction kits, has become (and is likely to continue to be) a major bottleneck as the pandemic expands.

Of particular importance is the ability to survey large asymptomatic populations- (1) to trace asymptomatic COVID-19 carriers which are otherwise difficult to identify and isolate; (2) to assure key personnel (e.g. healthcare personnel) are not contagious; (3) to screen high risk populations (such as nursing homes) to help protect them; (4) to accurately estimate the spread of the infection and the effectiveness of community measures and social distancing; and (5) to allow and monitor a safe return to work. Clearly, more efficient and higher-throughput diagnostic approaches are needed to support such efforts.

Several attempts to address this challenge have already been suggested, that can be categorized into three major approaches. The first approach is to replace PCR based methods by other direct diagnostic methods such as Loop-mediated Isothermal Amplification (LAMP)^1–3^ and CRISPR based diagnostic tools^4–6^, the second approach involves serological surveys^7–10^, and the third approach involves the improvement of the PCR methods capacity by optimization and automation^11–13^ or by reducing the required number of tests via pooling samples together, known as group testing.

Group testing is a field of research in the intersection of mathematics, computer science and information theory, with applications in biology, communication and more. A group testing algorithm is a testing scheme which is directed towards minimizing the number of tests conducted on a set of samples by using the ability to test pooled subsets of samples. If a pool of *n* samples tests negative, all samples must be negative, and therefore their status has been determined in only one test instead of *n* individual tests. Various group testing algorithms exist, with different assumptions and constraints^14,15^. While many such algorithms, most notably binary splitting, may be very efficient in theory, they might be unsuitable because of practical limitations. Three such limitations might be: (1) a limit on the number of stages due to the importance of delivering a test result quickly, exemplified by the urgent clinical context of COVID-19 diagnosis; (2) a limit on the ability to dilute samples and still safely identify a single positive sample in a pool; (3) favorability of simple algorithms which may minimize human error in a laboratory setting.

While several pooling approaches for SARS-CoV-2 detection were recently suggested^16–21^, these works mostly discussed theoretical considerations. Here we describe and demonstrate practical pooling solutions that save time and reagents by performing RNA extraction and RT-PCR on pooled samples. Unlike other suggestions of large-scale pooling and non-PCR-based methods, we maintain high sensitivity, and therefore the method complies with current clinical requirements. The simplicity of the method, similarity to currently approved procedures, and the fact that we do not require special sample handling or additional information make it easily adoptable on a large scale. We offer two such pooling approaches, based either on simple (Dorfman) pooling or matrix pooling^22,23^, and demonstrate their efficiency and sensitivity in the daily reality of SARS-CoV-2-infected clinical cases. Under current clinical diagnosis parameters these methods allow 5-fold to 7.5-fold increase in throughput when applied to populations with < 1% positives, including screened asymptomatic healthcare personnel and essential industries’ employees.

## Results

At the Hadassah Medical Center, two distinct populations of people are tested for SARS-CoV-2 at present. First, we receive samples from symptomatic patients, from the hospital and from the community. In these samples, about 10% of SARS-CoV-2 tests are positive. Second, we receive samples from prospectively screened asymptomatic populations such as hospital employees and workers in essential industries. Among the latter, individual testing of >2,000 samples revealed that the rate of positives ranged between 0.1% and 1%. Based on these findings, we examined the feasibility of pooled testing for asymptomatic populations, and designed appropriate pooling strategies.

A key requirement of pooled RNA extraction and RT-PCR tests is to retain sufficient sensitivity. In our RT-PCR assay, a sample is defined as *positive* if the viral genome is detected at threshold cycle (Ct) values ≤35, as *indeterminate* at Ct values >35 and ≤38, and as *negative* at Ct values >38. Theoretically, pooling 8 samples should elevate the Ct of a single positive sample by 3 cycles, and pooling 16 samples should elevate the Ct by 4 cycles. However, reproducibility of RNA extraction and RT-PCR might be affected by other factors. We therefore empirically tested the assay sensitivity, when multiple negative samples and one positive sample were mixed at the lysate stage. As shown in **Figure 1**, positive samples were readily detected, even when their individual Ct ranged between 35 and 38. Thus, SARS-CoV-2 RNA can be reliably detected in pooled samples without compromising the assay sensitivity.

**Figure 1:**
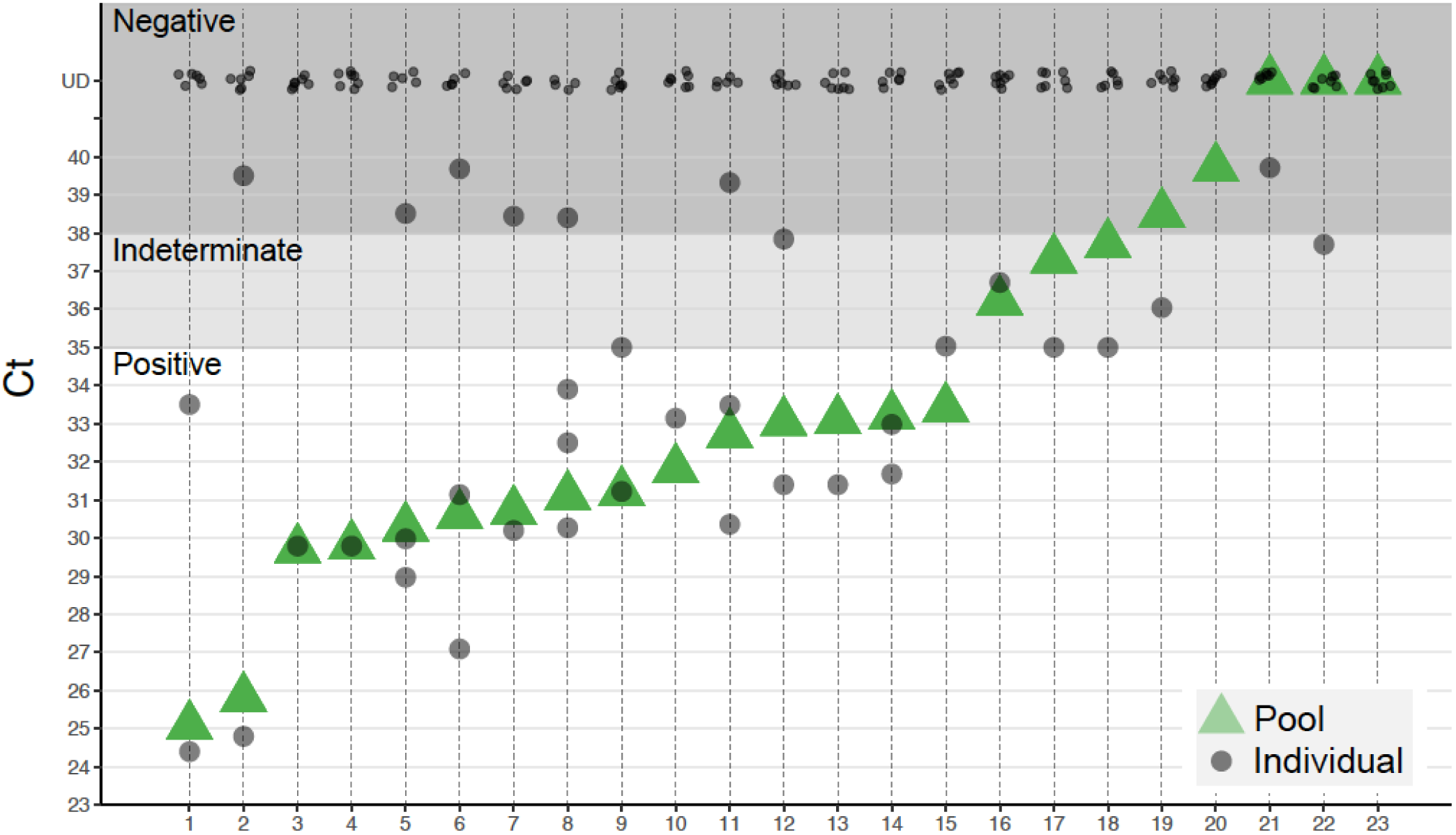
Pooling 8 lysates retain clinical sensitivity. Shown are results of 23 pooling experiments, with 8 lysates in each pool; 15 pools with positive samples indeed come up positive (pools 1–15), 3 pools without positive samples come up negative (pools 20, 21, 23), and 4 out of 5 pools containing a single indeterminate sample detected as indeterminate (pools 16, 17, 18, 19, 22); Pools containing 1–2 samples with low amount of SARS-CoV-2 are detected at a similar Ct (pools 9–18), showing clinical sensitivity is retained and the risk of false negatives is minimal. UD= Undetected.

The first pooling strategy is a simple two-stage testing algorithm known as Dorfman pooling^24^. In the first stage, the samples are divided into disjoint pools of *n* samples each, and each such pool is tested. A negative result implies that all samples in the pool are negative, while a positive result implies that at least one sample in the pool is positive. In the second stage, the samples of each pool that tested positive are individually tested. While such approaches for SARS-CoV-2 have been recently suggested, we have tested direct pooling of lysates of clinical nasopharyngeal samples, with RNA extraction already performed on the pooled samples. First, we tested the pooling of 184 consecutive samples into 23 pools of 8 samples each, and also tested in parallel each sample individually. This approach yielded highly accurate results, with no loss of diagnostic assay sensitivity: each of the pools that contained one or more positive samples was found to be positive, and all the pools that contained only negative samples were found to be negative (**Figure 1**). Of the 5 pools which contained one individual sample with an “indeterminate” result (in each pool), one was found to be negative suggesting potential yet negligible loss of sensitivity.

To further reduce the need to retest positive pools we have also tested a two-stage matrix pooling strategy^22,23^, where *n^2^* samples are ordered in an *n × n* matrix. Each row and each column are pooled, resulting in *2n* tests, 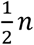 times less tests than individual testing. If either the number of positive rows or columns is one, the positive samples can be uniquely identified at the intersections of the positive rows and columns. Otherwise, if both the number of positive rows and columns is greater than one, intersections of positive rows and columns will be retested individually. We have tested this approach with pooling 75 samples into three *5 × 5* matrices, and identified all positive samples accurately (**Figure 2**). Importantly, the positive samples were detected in both the row and the column pools at a similar cycle in all three tested matrices, suggesting the pooling scheme is robust.

**Figure 2:**
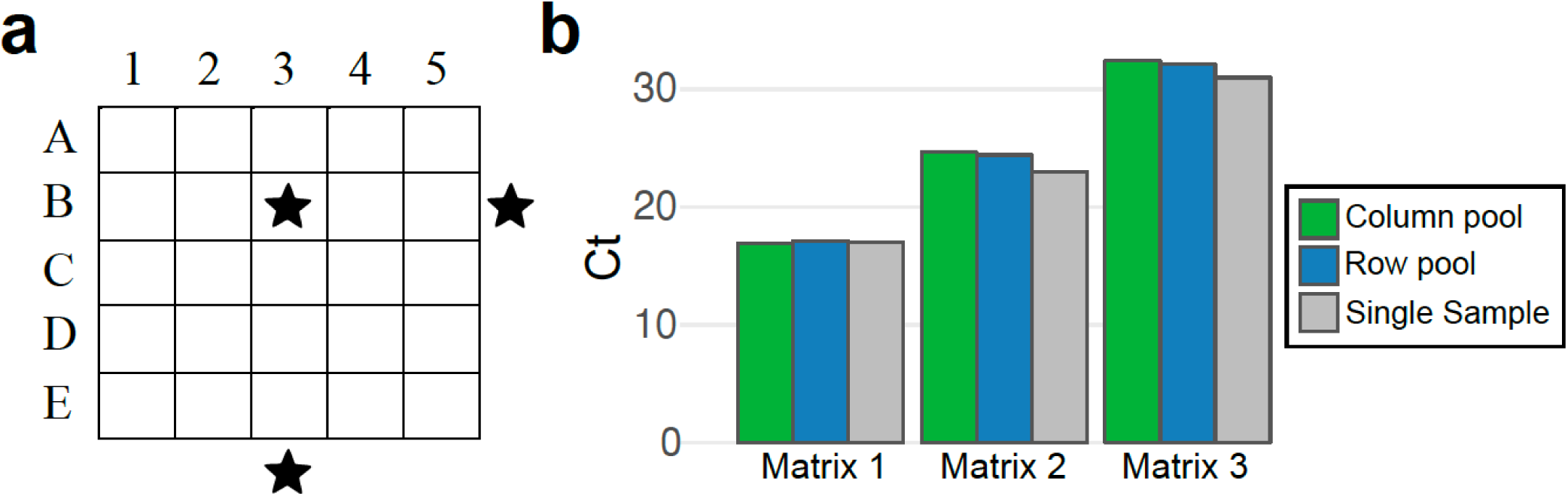
Matrix pooling. (**a**) Scheme for 5×5 matrix pooling. 25 samples sorted in a 5 × 5 matrix and each row and each column is pooled into a total of 10 pools, on which RNA extraction, reverse transcription and qPCR are performed. In this illustration row B and column 3 are positive (black stars), hence sample B3 is the only positive sample. If more than one row and one column are positive then all the samples in the intersection need to be retested, as some may be negative. (**b**) Three 5×5 pool matrices were generated (30 pools from 75 lysates). Each matrix (25 lysates that were previously tested individually) included a single lysate positive for SARS-COV-2. As expected, only 6 pools (one row and one column per matrix) were positive for SARS-COV-2, while 24 pools had Ct >40 (Undetected). RT-PCR Ct values of positive pools were nearly identical in the column pool (green) and the row pool (blue), and similar to the values of the individual test of the positive sample (gray).

Given the successful validation of both pooling strategies, we have adopted a Dorfman pooling protocol of 1:8 and employed it for the routine testing of nasopharyngeal swab samples from screened asymptomatic healthcare personnel, employees of essential industries, and residents and employees of nursing homes. Individual barcoded samples were received at the laboratory, inactivated by lysis buffer, pooled on a Tecan liquid-handling robot, and the pools were processed on a Qiasymphony robot for RNA extraction, and analyzed by RT-PCR. Results were interpreted and samples in positive pools were subsequently individually tested. Note that analysis of pool results requires close attention to indeterminate-result pools, showing a signal at 35 < Ct < 38, as these may contain individual positive samples. Therefore, criteria for retesting pools must be more stringent, e.g. a signal with Ct ~38 would be defined as negative when individual samples are tested, but warrants re-testing of individual samples when encountered in a pool (see **Table 1**, batch 3).

**Table 1:** Summary of pooled tests run at the Hadassah Medical Center. Ct-cycle threshold. * According to the hospital’s protocol for indeterminate values, RT-PCR was repeated with a different kit, and eventually was determined as positive.

|  | No. of samples | No. of pools | No. of positive pools | Ct of positive pools | No. of <u>positive</u> individual samples in the positive pools | Ct of positive individual samples | No. of total tests | No. of tests saved, compared to single sample tests (%) |
| --- | --- | --- | --- | --- | --- | --- | --- | --- |
| Pool batch 1 | 720 | 90 | 1 | 21.8 | 1 | 19.4 | 98 | 622 (86.4%) |
| Pool batch 2 | 720 | 90 | 0 | - | - | - | 90 | 630 (87.5%) |
| Pool batch 3 | 728 | 91 | 3 positives, 1 indeterminate | 25.39 | 2 | 22.05, 28.72 | 123 | 605 (83.1%) |
|  |  |  |  | 37.44 | 1 | 37.67* |  |  |
|  |  |  |  | 35.26 | 1 | 34.66 |  |  |
|  |  |  |  | 38.67 indeterminate | 0 | 38.24 (determined as negative) |  |  |
| Pool batch 4 | 3296 | 412 | 1 | - | 1 | 19 | 420 | 2876 (87.3%) |

At the time of submission of this manuscript, we have already tested 5,464 samples by pooling, thereby using 731 RNA extraction and RT-PCR reactions (a mere 13% of kits that would have been used in the full individual testing). Among these samples, we have identified and individually validated six positive samples, corresponding to a rate of 0.11% (**Table 1**).

## Discussion

An important consideration before implementing group testing is the expected rate of false positive and false negative results. Based on our experience with >2000 samples from asymptomatic individuals, we did not encounter any false positives in the pools, as shown in **Figure 1** and **Table 1**. False negatives are in principle more worrisome when testing in pools, because samples that failed at the RNA extraction step will be missed (while our individual testing includes amplification of a human transcript serving as an internal control for proper RNA extraction and RT-PCR of each sample). To define the magnitude of this potential problem, we examined a set of 13,781 tests done at our center, which were all expected to show a signal for a human gene serving as internal assay control. Amplification of the human gene failed in 52 samples (0.38% of the cases). Thus, we estimate that our current protocol of pooled sampling carries a risk of missing 0.38% of the positive samples. In a population of 1,000,000 individuals tested, of which 1,000 are positive (rate of 0.1%), this predicts that 4 positive individuals will be missed when using pools. We posit that this is a tolerable situation, particularly given the potentially much higher rates of false negative results due to swab sampling and other errors upstream.

The prevalence of COVID-19 in the tested population is not always known, which could affect the optimal pool size. This could be addressed either by other external estimates, such as a previous run of individual samples, rate of symptomatic patients, or alternative methods such as serological screening or wastewater titers monitoring^25^. Alternatively, it is possible to dynamically adapt pooling sizes, when the measured rate of positive samples is different than expected. Finally, there exist some group testing algorithms^15,26^ for the purpose of estimating the number of positive samples while using a relatively small (logarithmic) amount of tests, and such algorithms may be adapted to clinical constraints and parameters.

Future improvement of the sensitivity of the test, such as better sets of primers, improved sample collection and inclusion of information about pre-test probability will allow retaining sensitivity even when pooling a large number of sample lysates together. This will enable further improving efficiency, especially when prevalence is low, by increasing the pool size.

In summary, we demonstrate in a real-life situation the usefulness of pooled sampling starting at the early lysate stage. This saves time, work and reagents, allowing a considerable throughput increase of clinical diagnostic labs and opening the door for efficient screening of large asymptomatic populations for the presence of SARS-CoV-2 infection. Specifically, we have demonstrated that pooling lysates from 5 or 8 nasopharyngeal swab samples retains the sensitivity of viral RNA detection, allowing identification of SARS-CoV-2-positive individuals, and expected throughput increase of 5-fold to 7.5-fold.

## Methods

### Study Approval

These studies were part of the approved diagnostic procedures and optimization at the Hadassah Medical Center.

### Sample collection, RNA extraction and RT-PCR Detection

Nasopharyngeal swab samples were collected in 2 ml Viral Transport Medium (VTM) and mixed 1:1 with a 2x concentrated Zymo lysis buffer, or collected directly to 2 ml Zymo lysis buffer. For the initial validation, samples were collected from symptomatic patients or from screened healthy asymptomatic subjects. For sample lysate preparation 220 µL of sample VTM were added to 280 µL lysis buffer. RNA was extracted using MagNA Pure 96 kit (Roche Lifesciences) using Roche platform and eluted in 60 µL. 10 µL of RNA was used in 30 µL reaction using Real-Time Fluorescent RT-PCR kit (BGI).

### Pool RNA extraction and RT-PCR Detection

For matrix pool design we pooled equal volumes of sample lysate from each of the subjects to a final volume of 450 µL and used MagNA Pure 96 kit (Roche Lifesciences) using Roche platform. As supply was limited for this kit, we have used QIAsymphony DSP Virus/Pathogen kit on Qiasymphony platform for 1:8 pool design. We pooled equal volumes of sample lysate from each of the subjects to a final volume of 400 µL. Positive 1:8 pools were validated by single tests using QIAsymphony RNA kit on Qiasymphony platform. Both Qiagen kits were used with Zymo lysis buffer, and therefore we skipped the lysis and Proteinase K step. RNA was eluted into 60 µL; 10 µL of RNA was used for a 30 µL reaction using Real-Time Fluorescent RT-PCR kit (BGI).

### Choosing pooling strategy and parameters

We define the *efficiency* of a pooling algorithm as the total number of samples divided by the expected number of tests conducted on them. We assume all samples are independent and identically distributed, and denote the probability of a sample to be positive by *p* (prevalence of detectable COVID-19 patients in the relevant population) and the pool size by *n*. The efficiency of the algorithms described above depends on both *p* and *n*. The best theoretical efficiency is (−*plog*_2_(*p*)−(1−*p*)*log*_2_(1−*p*))^−1^ ^27^. The efficiency of Dorfman pooling is 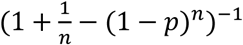^24^. We chose a pool size of *n=8* samples as it allows low false negative rate (**Figure 1**) and high efficiency for a wide range of COVID-19 prevalence (**Table 2**). The prevalence of detectable COVID-19 in an asymptomatic population is estimated to be considerably below 1%^28^, and indeed of the 5,464 asymptomatic subjects tested in the present study only 0.11% were found positive. Therefore, efficiency is likely to be 5 - 7.5. For higher prevalence the efficiency of matrix pooling is somewhat higher (see **Table 2** and supplementary note). We provide a tool (https://github.com/matanseidel/pooling_optimization) to help choose the approach and pool size based on the prevalence. Even when matrix pooling is not more efficient, it may have other benefits, as it significantly reduces the need for retesting, and provides additional confidence that a sample is negative as it is detected as negative in two pools.

**Table 2:** Efficiency of Dorfman and matrix pooling with pool size n=8 compared with optimal efficiency.

| Prevalence ($p$) | 0.1% | 1% | 2% | 5% | 10% | 20% |
| --- | --- | --- | --- | --- | --- | --- |
| Maximal theoretical efficiency | 87.7 | 12.4 | 7.1 | 3.5 | 2.13 | 1.39 |
| Dorfman $n=8$ efficiency | 7.5 | 4.9 | 3.6 | 2.2 | 1.44 | 1.04 |
| Matrix $n=8$ efficiency | 4.0 | 3.9 | 3.6 | 2.6 | 1.68 | 1.05 |

In the case that samples are not independent, and we have information regarding their dependency, we can try to further efficiency by grouping together dependent samples, that is, samples that are likely all positives or all negatives, such as members of the same family, or samples that are likely to be all negative since they have a low risk profile. This will increase the number of negative pools, and therefore decrease the overall number of tests conducted.

## Data Availability

All data appear in the main text.

## Acknowledgments

Yotam Drier is supported by an Alon Fellowship and the Concern Foundation / American Friends of the Hebrew University Young Professorship Award. The following people are members of the Hebrew University-Hadassah Medical School COVID-19 diagnosis team and have contributed to the development of the pooled testing pipeline: Agnes Klochendler, Amir Eden, Avihu Klar, Avraham Geldman, Ayelet Arbel, Ayelet Peretz, Batel Shalom, Bracha Lea Ochana, Dana Avrahami-Tzfati, Daniel Neiman, Daniel Steinberg, Danny Ben Zvi, Etai shpigel, Gal Atlan, Hagar Klein, Hanna Chekroun, Haran Shani, Idit Hazan, Ihab Ansari, Itia Magenheim, Joshua Moss, Judith Magenheim, Liat Peretz, Libi Feigin, Malkiel Saraby, Maya Sherman, Mercedes Bentata, Mevaseret Avital, Miriam Kott, Moriah Peyser, Moriya Weitz, Moriyah Shacham, Myriam Grunewald, Naama Sasson, Nadav Wallis, Narmen Azazmeh, Netanel Tzarum, Ori Fridlich, Raz Sher, Reba Condiotti, Ron Refaeli, Roni Ben Ami, Roni Zaken Gallili, Ronny Helman, Shai Ofek, Shay Tzaban, Sheina Piyanzin, Shira Anzi, Shira Dagan, Sivan Lilenthal, Tal Sido, Tamar Licht, Tomer Friehmann, Yael Kaufman, Amir Pery, Ann Saada, Assaf Dekel, Avner Yeffet, Avraham Shaag, Ayelet Michael Gayego, Bracha-Lea Ochana, Ela Shay, Eliran Arbib, Hadil Onallah, Kerem Ben-Meir, Leonid Levinzon, Leonor Cohen-Daniel, Luba Natan, Marah Hamdan, Mila Rivkin, Mohammad Shwieki, Olesya Vorontsov, Rimma Barsuk, Rinat Abramovitch, Rita Gutorov, Salem Sirhan, Suhair Abdeen, Yulia Yachnin, Yutti Daitch.

## References

1. Zhang, Y. et al. Rapid Molecular Detection of SARS-CoV-2 (COVID-19) Virus RNA Using Colorimetric LAMP. Infectious Diseases (except HIV/AIDS) (2020) doi:10.1101/2020.02.26.20028373.

2. Yan, C. et al. Rapid and visual detection of 2019 novel coronavirus (SARS-CoV-2) by a reverse transcription loop-mediated isothermal amplification assay. Clin. Microbiol. Infect. (2020) doi:10.1016/j.cmi.2020.04.001.

3. Park, G.-S. et al. Development of Reverse Transcription Loop-mediated Isothermal Amplification (RT-LAMP) Assays Targeting SARS-CoV-2. J. Mol. Diagn. (2020) doi:10.1016/j.jmoldx.2020.03.006.

4. Ai, J.-W., Zhang, Y., Zhang, H.-C., Xu, T. & Zhang, W.-H. Era of molecular diagnosis for pathogen identification of unexplained pneumonia, lessons to be learned. Emerg. Microbes Infect. 9, 597–600 (2020).

5. Lucia, C., Federico, P.-B. & Alejandra, G. C. An ultrasensitive, rapid, and portable coronavirus SARS-CoV-2 sequence detection method based on CRISPR-Cas12. bioRxiv 2020.02.29.971127 (2020) doi:10.1101/2020.02.29.971127.

6. Broughton, J. P. et al. Rapid Detection of 2019 Novel Coronavirus SARS-CoV-2 Using a CRISPR-based DETECTR Lateral Flow Assay. Infectious Diseases (except HIV/AIDS) (2020) doi:10.1101/2020.03.06.20032334.

7. Guo, L. et al. Profiling Early Humoral Response to Diagnose Novel Coronavirus Disease (COVID-19). Clin. Infect. Dis. (2020) doi:10.1093/cid/ciaa310.

8. Xiao, S.-Y., Wu, Y. & Liu, H. Evolving status of the 2019 novel coronavirus infection: Proposal of conventional serologic assays for disease diagnosis and infection monitoring. Journal of medical virology vol. 92 464–467 (2020).

9. Zhang, W. et al. Molecular and serological investigation of 2019-nCoV infected patients: implication of multiple shedding routes. Emerg. Microbes Infect. 9, 386–389 (2020).

10. Lassaunière, R. et al. Evaluation of nine commercial SARS-CoV-2 immunoassays. Infectious Diseases (except HIV/AIDS) (2020) doi:10.1101/2020.04.09.20056325.

11. Fomsgaard, A. S. & Rosenstierne, M. W. An alternative workflow for molecular detection of SARS-CoV-2 - escape from the NA extraction kit-shortage. medRxiv 2020.03.27.20044495 (2020).

12. Barra, G. B., Santa Rita, T. H., Mesquita, P. G., Jacomo, R. H. & Nery, L. F. A. Analytical sensibility and specificity of two RT-qPCR protocols for SARS-CoV-2 detection performed in an automated workflow. Infectious Diseases (except HIV/AIDS) (2020) doi:10.1101/2020.03.07.20032326.

13. Kalikiri, M. K. R. et al. High-throughput extraction of SARS-CoV-2 RNA from nasopharyngeal swabs using solid-phase reverse immobilization beads. Infectious Diseases (except HIV/AIDS) (2020) doi:10.1101/2020.04.08.20055731.

14. Du, D., Hwang, F. K. & Hwang, F. Combinatorial Group Testing and Its Applications. (World Scientific, 2000).

15. Aldridge, M., Johnson, O. & Scarlett, J. Group Testing: An Information Theory Perspective. Foundations and Trends® in Communications and Information Theory 15, 196–392 (2019).

16. Yelin, I. et al. Evaluation of COVID-19 RT-qPCR test in multi-sample pools. Infectious Diseases (except HIV/AIDS) (2020) doi:10.1101/2020.03.26.20039438.

17. Sinnott-Armstrong, N., Klein, D. & Hickey, B. Evaluation of Group Testing for SARS-CoV-2 RNA. Infectious Diseases (except HIV/AIDS) (2020) doi:10.1101/2020.03.27.20043968.

18. Shani-Narkiss, H., Gilday, O. D., Yayon, N. & Landau, I. D. Efficient and Practical Sample Pooling High-Throughput PCR Diagnosis of COVID-19. Public and Global Health (2020) doi:10.1101/2020.04.06.20052159.

19. Deckert, A., Bärnighausen, T. & Kyei, N. Pooled-sample analysis strategies for COVID-19 mass testing: a simulation study. nCoV (2020).

20. Shental, N. et al. Efficient high throughput SARS-CoV-2 testing to detect asymptomatic carriers. Infectious Diseases (except HIV/AIDS) (2020) doi:10.1101/2020.04.14.20064618.

21. Gollier, C. & Gossner, O. Group Testing Against Covid-19. EconPol Policy Brief 24, (2020).

22. Kwiatkowski, T. J., Jr, Zoghbi, H. Y., Ledbetter, S. A., Ellison, K. A. & Chinault, A. C. Rapid identification of yeast artificial chromosome clones by matrix pooling and crude lysate PCR. Nucleic Acids Res. 18, 7191–7192 (1990).

23. Barillot, E., Lacroix, B. & Cohen, D. Theoretical analysis of library screening using a N-dimensional pooling strategy. Nucleic Acids Res. 19, 6241–6247 (1991).

24. Dorfman, R. The Detection of Defective Members of Large Populations. Ann. Math. Stat. 14, 436–440 (1943).

25. Wu, F. et al. SARS-CoV-2 titers in wastewater are higher than expected from clinically confirmed cases. medRxiv 2020.04.05.20051540 (2020).

26. Damaschke, P. & Sheikh Muhammad, A. Competitive Group Testing and Learning Hidden Vertex Covers with Minimum Adaptivity. in Fundamentals of Computation Theory 84–95 (Springer Berlin Heidelberg, 2009).

27. Li, T., Chan, C. L., Huang, W., Kaced, T. & Jaggi, S. Group testing with prior statistics. in 2014 IEEE International Symposium on Information Theory 2346–2350 (2014).

28. Lavezzo, E. et al. Suppression of COVID-19 outbreak in the municipality of Vo, Italy. Epidemiology (2020) doi:10.1101/2020.04.17.20053157.

